# Pathologic Response to Immunotherapy is Associated with Survival in Patients Undergoing Delayed Nephrectomy for Metastatic Renal Cell Carcinoma

**DOI:** 10.1101/2025.04.24.25326353

**Authors:** Yash S. Khandwala, Stephen W. Reese, Burcin A. Ucpinar, Andrea Knezevic, Lennert Eismann, Chih-Ying Wu, Rohan Mittal, Sahil Doshi, Daniel Barbakoff, Andrea Lopez Sanmiguel, Jonathan Coleman, Mark Dawidek, Robert J. Motzer, Paul Russo, Oguz Akin, Ritesh R. Kotecha, Yingbei Chen, Martin H. Voss, A. Ari Hakimi

**Author notes:** Corresponding Author: A. Ari Hakimi, MD, Department of Surgery, Urology Service 1275 York Avenue, New York, NY 10065, Memorial Sloan Kettering Cancer Center.

## Abstract

**Introduction:** The role of consolidative nephrectomy (CN) in metastatic renal cell carcinoma (mRCC) patients treated with immune checkpoint inhibitors (ICIs) remains unknown. As patients derive variable benefit from immunotherapy (IO), understanding how treatment response correlates with long-term outcomes could inform patient surgical selection. We thus conducted a retrospective study to evaluate whether radiographic and pathologic tumor responses after immunotherapy are associated with survival in patients undergoing CN at a high-volume academic center.

**Methods:** We identified mRCC patients treated with at least one IO-containing regimen followed by CN between 2015 and 2024. Radiographic response was assessed using RECIST. Pathologic response was measured using percent residual viable tumor (RVT), with major pathologic response (MPR) defined as RVT <10%. Outcomes included progression-free (PFS) and overall survival (OS), analyzed using Cox proportional hazards models.

**Results:** Sixty patients underwent CN after immunotherapy. Median time to nephrectomy was 9 months (IQR: 7, 14) and 2-year OS was 75% (95% CI: 60-85). Fifteen patients (26%) had an MPR and 21 patients (35%) had a radiographic response of the primary tumor. Radiographic response was moderately correlated with RVT (Spearman correlation 0.51, p <0.001) and was protective but not significantly associated with PFS or OS. MPR was significantly associated with PFS (HR: 0.05, 95% CI: 0.01-0.41; p=0.005) and OS (HR: 0.07, 95% CI: 0.01-0.88; p=0.04).

**Conclusion:** Patients with MPR at nephrectomy had longer PFS and OS. Pathologic response may help guide post-nephrectomy treatment timing and sequencing, though future efforts should further validate the utility of post-IO pathologic endpoints.

## Introduction

Surgical management of metastatic renal cell carcinoma (mRCC) has evolved over the past 30 years. During the cytokine era, upfront cytoreductive nephrectomy was an important mainstay of treatment and multiple clinical trials demonstrated an overall survival (OS) benefit compared to interferon alpha alone.^1–4^ However, with advances in systemic therapy, and with the results of two major clinical trials, CARMENA and SURTIME, upfront cytoreductive nephrectomy became reserved for select patients with low burden of disease and favorable risk factors who were most likely to benefit.^5–7^ SURTIME further solidified the paradigm of administering systemic therapy prior to consolidative surgical therapy by demonstrating a higher median overall survival with this sequence.^6^

More recently, immunotherapy (IO)-based doublet regimens have effectively replaced Tyrosine kinase inhibitor (TKI) monotherapy in the front-line metastatic setting, based on substantial primary and metastatic tumor responses seen in several modern clinical trials including CheckMate 214 (nivolumab and ipilimumab), KEYNOTE 426 (pembrolizumab and axitinib), and CLEAR (lenvatinib and pembrolizumab). This has prompted renewed interest in the role of CN in contemporary practice.^8–10^ A recent retrospective study found that delayed consolidative nephrectomy reduced the risk of death at 36 months by 55% compared to immunotherapy alone. They, along with other studies, also demonstrated that delayed nephrectomy can be performed safely with low rates of complications.^11–14^ However, there remains a lack of clinical guidance on when consolidation should be performed and what patient factors may help select candidates who would most benefit from surgery. Furthermore, little data exists on how surgical pathology should inform adjuvant treatment strategies in these select patients.

Thus, we performed a retrospective review of patients with mRCC undergoing consolidative nephrectomy after receiving IO-based therapy at our institution. We sought to define the clinical course and time frame in which these patients underwent consolidative nephrectomy and to identify whether radiographic and pathologic responses to ICI were associated with survival.

## Methods

### Study Population and Data Source

This was a retrospective cohort study including 60 mRCC patients who underwent consolidative nephrectomy after receiving an IO-containing therapy between April 2015 and June 2024. Using a prospectively maintained database of consecutive patients ≥ 18 years who underwent CN at our institution, we identified all patients who received at least one line of immunotherapy (inclusive of multiagent combinations) prior to surgery. The two indications for surgery were positive response to systemic therapy (consolidative-intent) and palliation for a symptomatic primary tumor (palliative-intent).

Patient and tumor characteristics, including International Metastatic RCC Database Consortium (IMDC) risk scores, were obtained from our institutional database which uses structured queries and natural language processing to extract clinicodemographic data.^15^ Comorbidities were captured using the Charlson Comorbidity Index (CCI) and scored as discrete count categories between 0 and 12.^16^ Primary and metastatic tumor size were measured using the sum of the longest diameters of target lesions as per standard RECIST criteria from either abdominal computed tomography (CT) or magnetic resonance imaging (MRI).^17^ Treatment data were obtained through manual electronic medical record (EMR) review.

### Outcomes

Patients were evaluated for progression-free (PFS) and overall survival (OS) after CN. Progression was defined by standard RECIST 1.1 criteria and OS was measured as time between CN and death.^17^ Patients without events were censored at last follow-up for both outcomes.

Patients were evaluated for pathologic response to therapy in the primary tumor at the time of nephrectomy. Pathologic response was measured using percent residual viable tumor (RVT) in the nephrectomy sample, which was calculated as residual viable tumor area / total tumor bed area (including viable tumor, necrosis, and regression tumor bed). The value was rendered by genitourinary pathologists (YBC, CYW, RM) by examining across all slides and incorporating information from gross examination. Major pathologic response to immunotherapy was defined as RVT < 10%, partial response as 10-50% RVT and no response as RVT > 50% based on standardized pathologic reporting guidelines published by the International Neoadjuvant Melanoma Consortium.^18^ Radiographic response was measured as the difference in longest diameter of the primary tumor from the initial scan to the scan immediately prior to surgery. We tested a radiographic response cut-off of at least 30% decrease in the longest primary tumor diameter which is loosely based off the RECIST 1.1 definition of a partial radiographic response being at least a 30% reduction from baseline sum of target lesions.^17^

### Statistical Analysis

The Kaplan-Meier estimator was used to estimate PFS and OS, and Cox proportional-hazards regression models were used to estimate the effect of radiographic and pathologic response on these outcomes after adjustment for surgical indication (palliation or consolidation) and number of metastatic sites at diagnosis. Associations between residual viable tumor and change in primary tumor diameter were assessed using Spearman’s correlation coefficient. Statistical analyses were conducted using SAS, version 9.4 (Cary, NC). All statistical tests were two-sided and p-values < 0.05 were considered statistically significant.

## RESULTS

Baseline characteristics of all 60 patients with metastatic RCC who underwent delayed nephrectomy after immunotherapy are presented in **Table 1**. Median age was 59 years (IQR: 52, 67); 53 (87%) had predominantly clear cell histology and 21 (35%) had sarcomatoid or rhabdoid features. The lung was the most common site of metastatic disease at presentation in 39 (74%) while 13 (24%) had liver metastases and 5 (9%) had brain metastases. Fifty patients (83%) underwent one line of therapy prior to surgery while the remainder of the cohort underwent a second line. Patients underwent upfront treatment with PD1/PDL1 + TKI inhibitors (32, 53%), CTLA4 + PD1 inhibitors (27, 45%) or PD-1 inhibitors alone (6, 10%). Median time from immunotherapy start to nephrectomy was 9 months (IQR: 7, 14). Post-CN systemic therapy data is presented in Supplementary Table 1. Thirty-one patients (52%) resumed the same regimen within 6 weeks after surgery while 16 (27%) were not restarted on any additional therapy. Medication re-start rates did not differ notably between patients who received CN for palliation, (8/17, 47%), and consolidation (23/43, 54%). Figure 1 presents the clinical course and treatment regimen of each patient.

**Figure 1.**
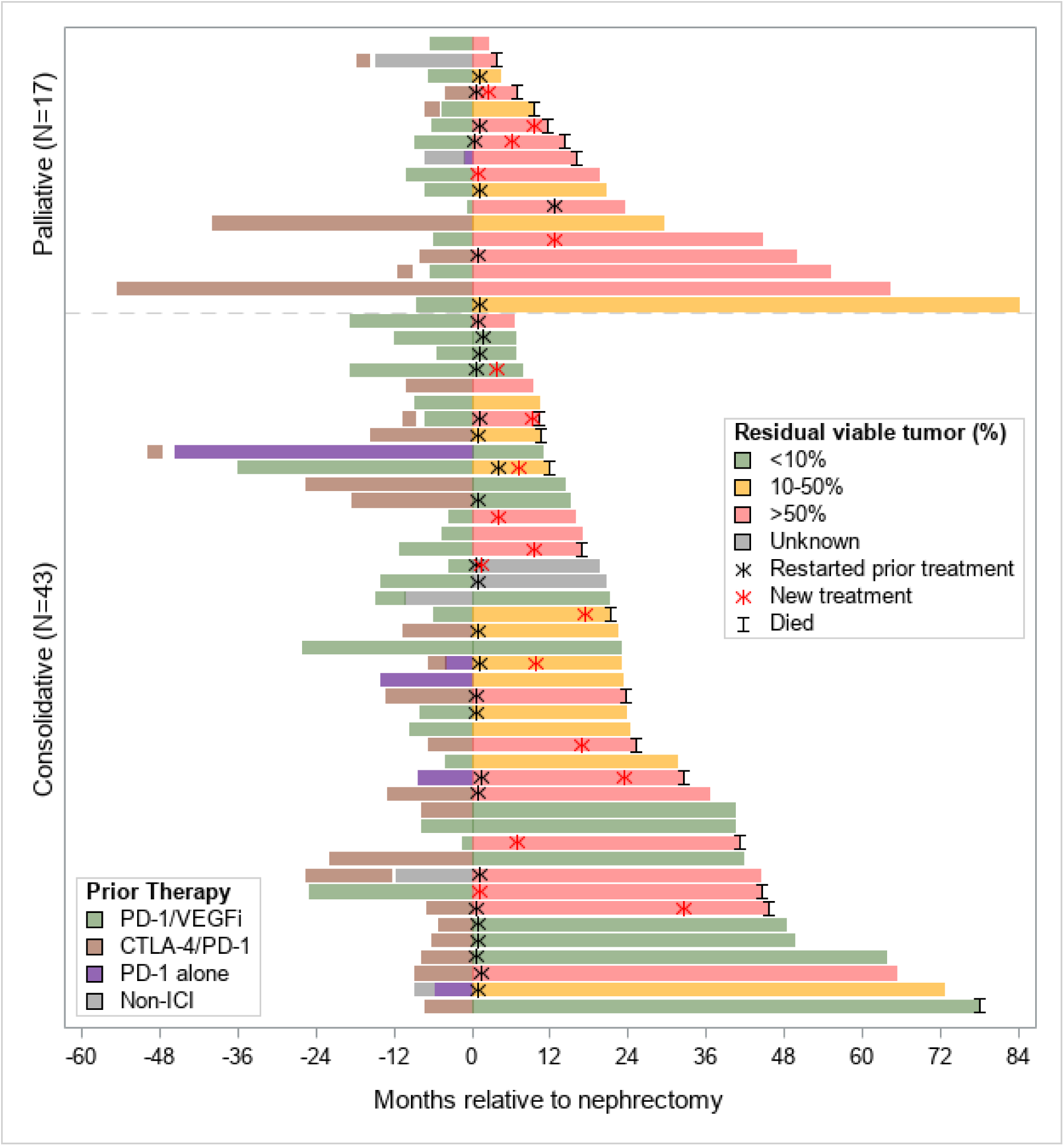
Swimmer plot depicting the duration of treatment course relative to nephrectomy date and clinical status at last follow-up of each subject.

**Table 1.** Patient characteristics and treatment (N=60)

|  | Frequency (%) or Median (IQR) |
| --- | --- |
| Age at nephrectomy, years | 59 (52, 67) |
| Sex |  |
| Male | 50 (83%) |
| Female | 10 (17%) |
| KPS <80* | 3 (6%) |
| Charlston comorbidity index† | 8 (8, 9) |
| Histology |  |
| Clear cell | 53 (87%) |
| Unclassified | 6 (10%) |
| Papillary | 2 (3%) |
| Sarcomatoid/rhabdoid features | 21 (35%) |
| Initial primary tumor diameter, cm† | 8.8 (7.1, 12.2) |
| Number of metastatic sites† | 3 (3, 4) |
| Metastatic sites† |  |
| Lung | 39 (74%) |
| Bone | 18 (34%) |
| Liver | 13 (24%) |
| Brain | 5 (9%) |
| IMDC risk* |  |
| Intermediate | 29 (57%) |
| Poor | 22 (43%) |
| Indication for nephrectomy |  |
| Consolidation | 43 (72%) |
| Palliation | 17 (28%) |
| Type of nephrectomy |  |
| Open | 33 (55%) |
| Robotic/Laparoscopic | 27 (45%) |
| Partial nephrectomy | 4 (7%) |
| Lines of therapy prior to nephrectomy |  |
| 1 | 50 (83%) |
| 2 | 10 (17%) |
| ICI regimen received |  |
| PD1/L1 + TKI | 32 (53%) |
| CTLA4 + PD1 | 27 (45%) |
| PD1 alone | 6 (10%) |
| Months from immunotherapy start to nephrectomy | 9 (7, 14) |

Abbreviations: KPS, Karnofsky performance score; IMDC, International mRCC database consortium; ICI, Immune checkpoint inhibitor; PD-1, Programmed cell death protein 1 inhibitor; VEGFi, Vascular endothelial growth factor inhibitor; CTLA-4, Cytotoxic T-Lymphocyte-Associated Protein 4 inhibitor.
\*Missing for 9 patients
†Missing for 7 patients

Patient outcomes for each surgical indication before and after nephrectomy are presented in **Table 2**. Median radiographic change of primary tumor diameter in response to therapy before nephrectomy was -21% (IQR: -37, 0). Median radiographic change of the sum of non-primary target lesions before nephrectomy was -54% (IQR: -77, -23). Fifteen patients (26%) had a major pathologic response with less than 10% RVT, including 9 (16%) with complete response (0% RVT) within the primary mass at the time of nephrectomy. MPR was seen after receipt of both CTLA4 + PD1 inhibitors (9, 16%), and PD1/PDL1 + TKI inhibitors (6, 10%). The association between change in primary tumor size and pathologic response is illustrated in **Figure 2**.

**Figure 2.**
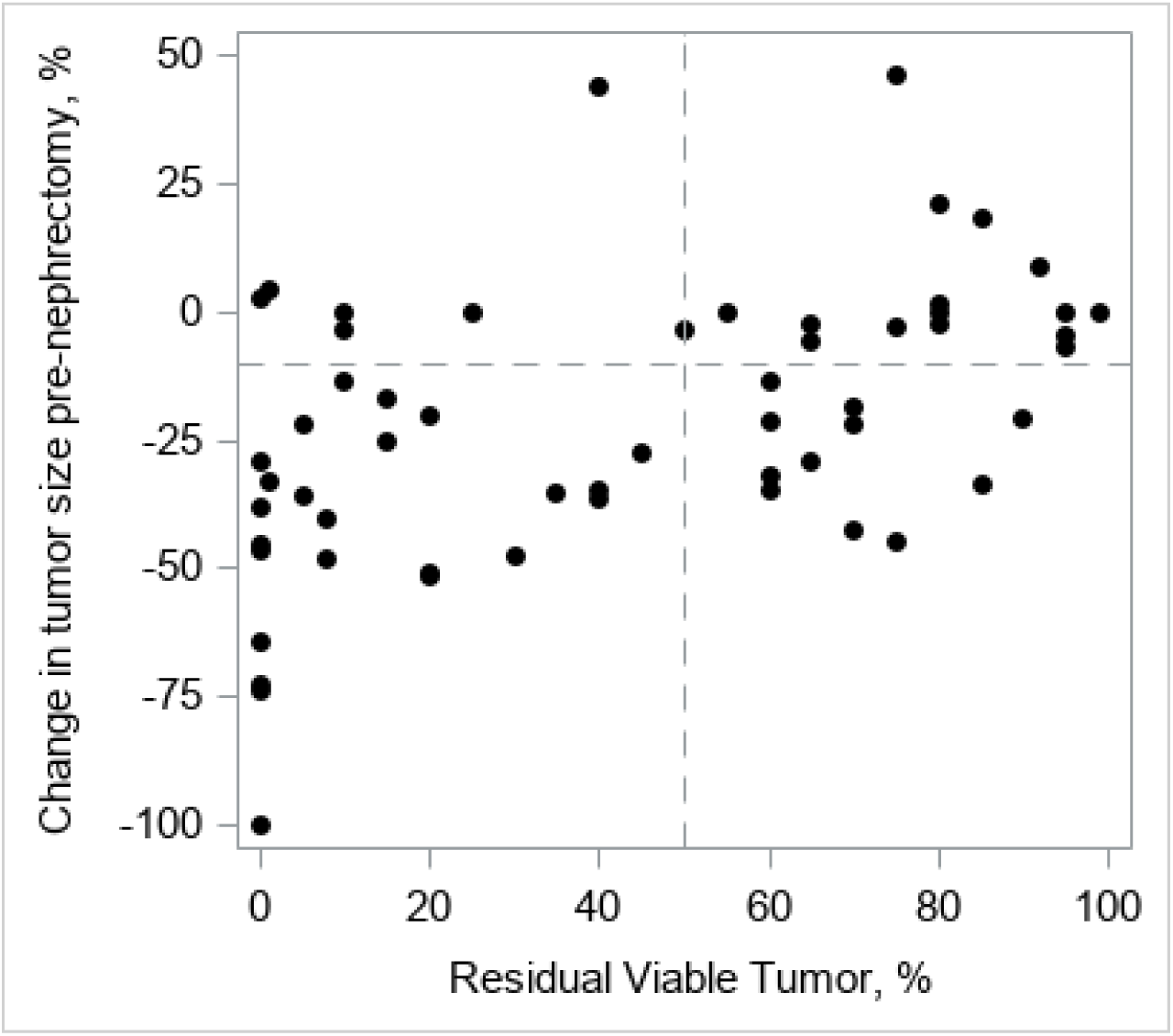
Change in radiographic primary tumor size (%) versus RVT (%). Reference lines defined by -10% change in radiographic primary tumor size and 50% RVT. A moderate positive correlation is observed between change in tumor size and residual viable tumor (Spearman correlation 0.51). In 40 patients with radiographic primary tumor shrinkage (less than -10% change in size from baseline), 24 had less than 50% RVT (lower-left quadrant; 2-year OS 95% (95% CI: 68, 99)) and 11 had greater than 50% RVT (lower-right quadrant; 2-year OS 78% (95% CI: 36, 94)).

**Table 2.**
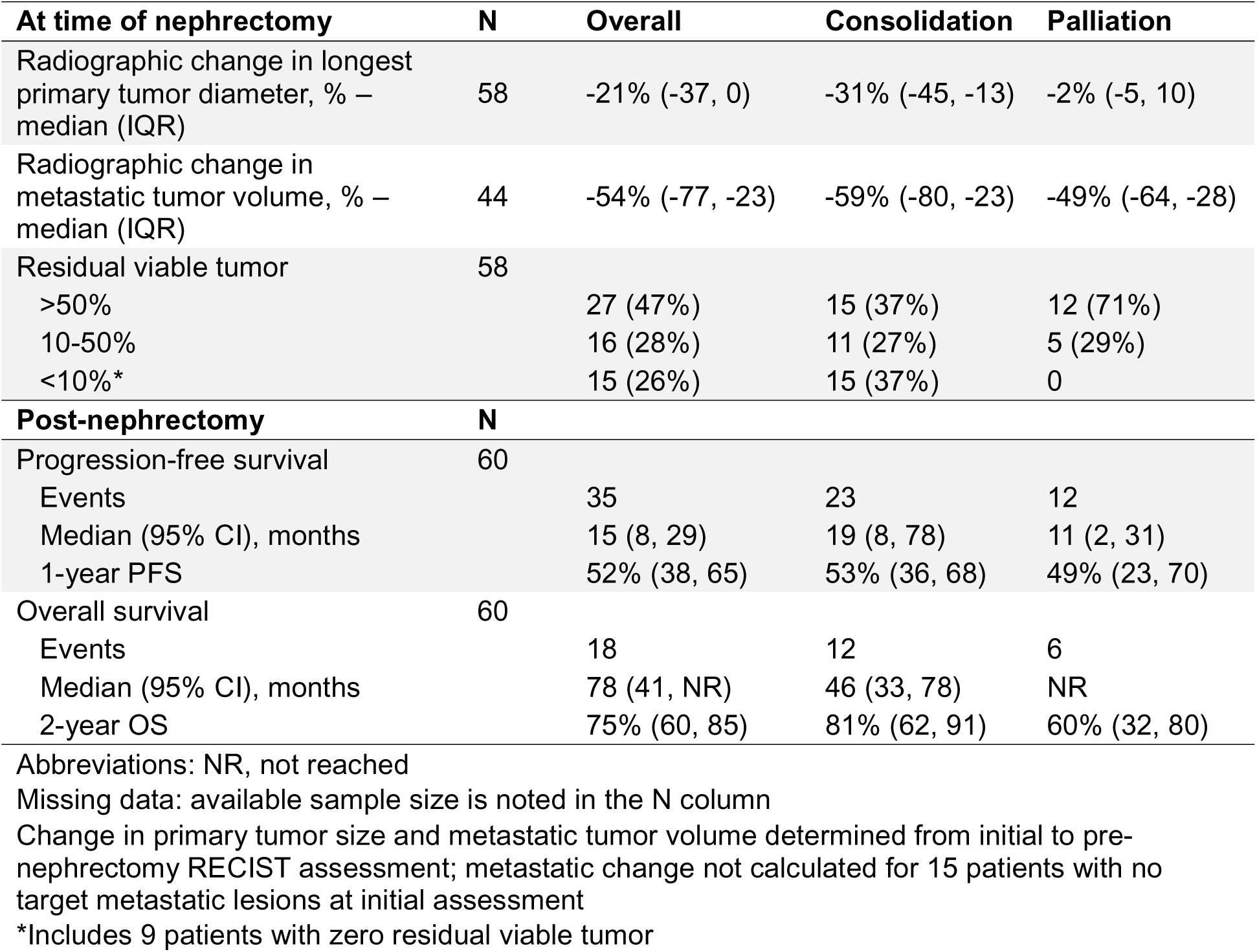
Outcomes by nephrectomy indication.

| **At time of nephrectomy** | **N** | **Overall** | **Consolidation** | **Palliation** |
| --- | --- | --- | --- | --- |
| Radiographic change in longest primary tumor diameter, % – median (IQR) | 58 | -21% (-37, 0) | -31% (-45, -13) | -2% (-5, 10) |
| Radiographic change in metastatic tumor volume, % – median (IQR) | 44 | -54% (-77, -23) | -59% (-80, -23) | -49% (-64, -28) |
| Residual viable tumor | 58 |  |  |  |
| >50% |  | 27 (47%) | 15 (37%) | 12 (71%) |
| 10-50% |  | 16 (28%) | 11 (27%) | 5 (29%) |
| <10%* |  | 15 (26%) | 15 (37%) | 0 |
| **Post-nephrectomy** | **N** |  |  |  |
| Progression-free survival | 60 |  |  |  |
| Events |  | 35 | 23 | 12 |
| Median (95% CI), months |  | 15 (8, 29) | 19 (8, 78) | 11 (2, 31) |
| 1-year PFS |  | 52% (38, 65) | 53% (36, 68) | 49% (23, 70) |
| Overall survival | 60 |  |  |  |
| Events |  | 18 | 12 | 6 |
| Median (95% CI), months |  | 78 (41, NR) | 46 (33, 78) | NR |
| 2-year OS |  | 75% (60, 85) | 81% (62, 91) | 60% (32, 80) |

Abbreviations: NR, not reached
Missing data: available sample size is noted in the N column
Change in primary tumor size and metastatic tumor volume determined from initial to pre-nephrectomy RECIST assessment; metastatic change not calculated for 15 patients with no target metastatic lesions at initial assessment
\*Includes 9 patients with zero residual viable tumor

Median PFS after nephrectomy was 15 months (95% CI: 8, 29) and 1-year PFS was 52% (95% CI: 38, 65). Median OS was 78 months (95% CI: 41, NR) and 2-year OS was 75% (95% CI: 60, 85). Two-year OS was 81% (95% CI: 62, 91) for patients undergoing CN for consolidation compared to 60% (95% CI: 32, 80) for palliation. Kaplan-Meier curves for PFS and OS of the cohort are presented in **Figure 3**.

**Figure 3a.**
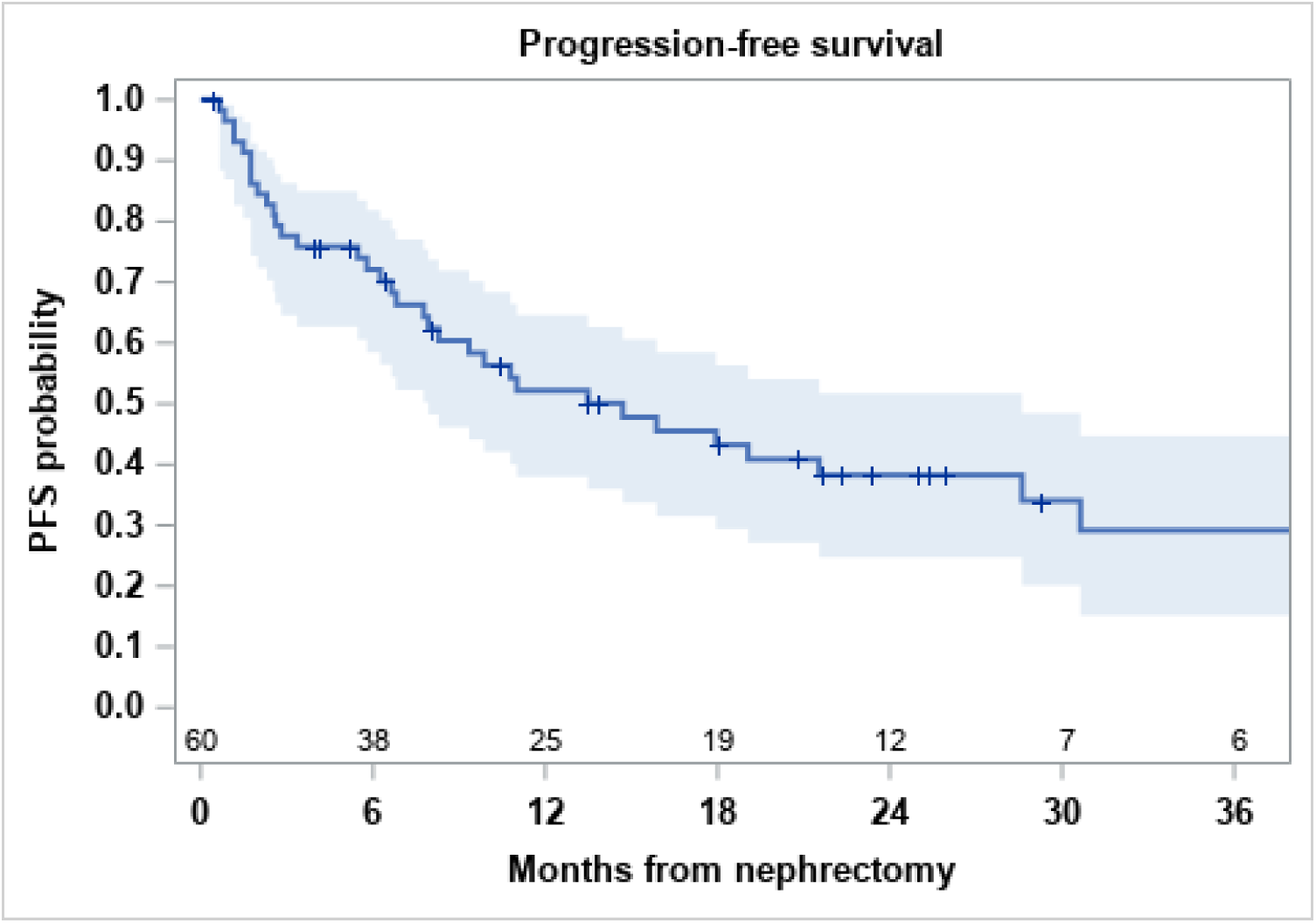
Progression-free survival post-nephrectomy. Of 60 patients, 34 experienced disease progression and 1 died with no documented progression (35 PFS events). Median PFS was 15 months (95% CI: 8 to 29) and 1-year PFS probability was 52% (95% CI: 38 to 65).

**Figure 3b.**
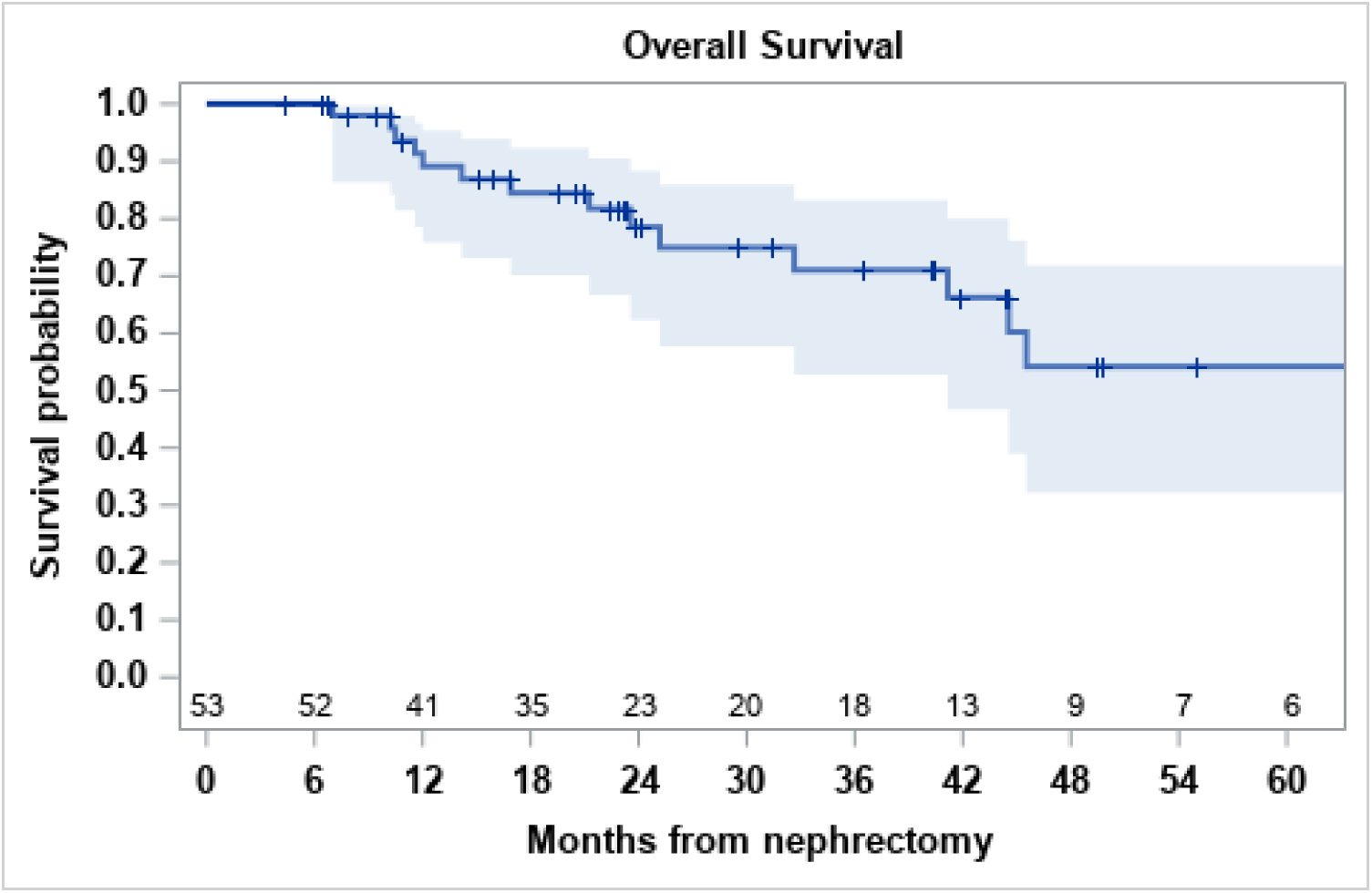
Overall survival post-nephrectomy. Of 53 patients with post-nephrectomy follow-up, death was observed in 18 patients while the 42 survivors had median follow-up time of 29 months (95% CI: 22 to 44). Median overall survival was 78 months (95% CI: 41 to NR) and 2-year survival probability was 75% (95% CI: 60 to 85).

A radiographic change in primary tumor size ≤ -30% was not significantly associated with improved PFS (HR: 0.45, 95% CI: 0.18,1.12; p=0.09) nor OS (HR: 0.70, CI: 0.17, 2.89; p=0.62).

We performed sensitivity analyses (not shown) at the -50% and -80% target levels which did not significantly alter the results. Major pathologic response of RVT < 10% was associated with both improved PFS (HR: 0.05, 95% CI: 0.01, 0.41; p=0.005) and OS (HR: 0.07, 95% CI: 0.01, 0.88; p=0.04) compared to RVT > 50% (**Table 3**). In patients with RVT < 10%, RVT 10-50%, and RVT > 50%, the 2-year OS was 100%, 76% (95% CI: 43, 92) and 71% (95% CI: 46, 86), respectively. Kaplan-Meier curves for PFS and OS by pathologic response are presented in **Figure 4**.

**Figure 4a.**
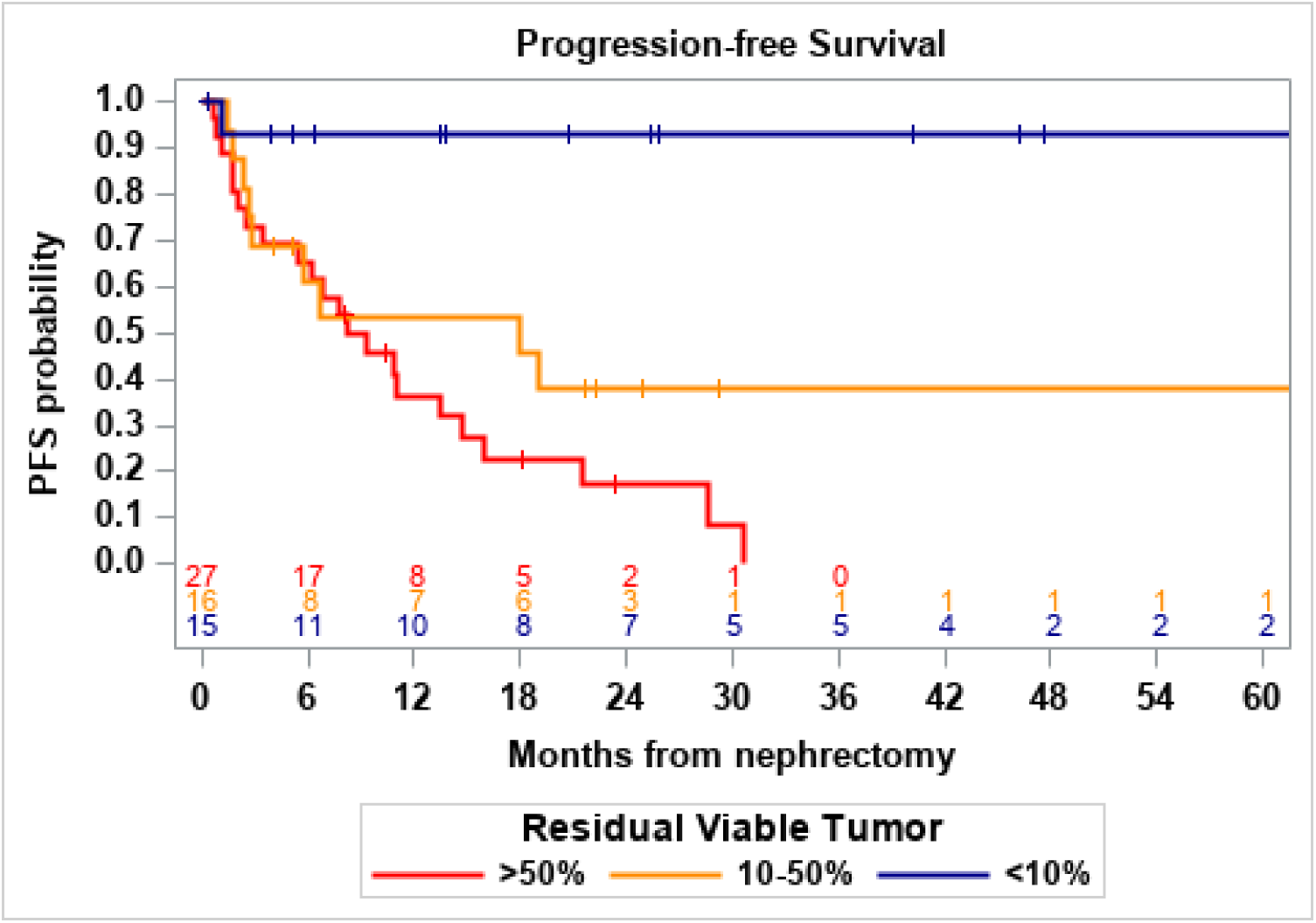
Progression-free survival by residual viable tumor. In patients with RVT <10%, 10-50% and >50%, 1-year PFS was 93% (95% CI: 59 to 99), 53% (95% CI: 26 to 75) and 36% (95% CI: 18 to 55), respectively.

**Figure 4b.**
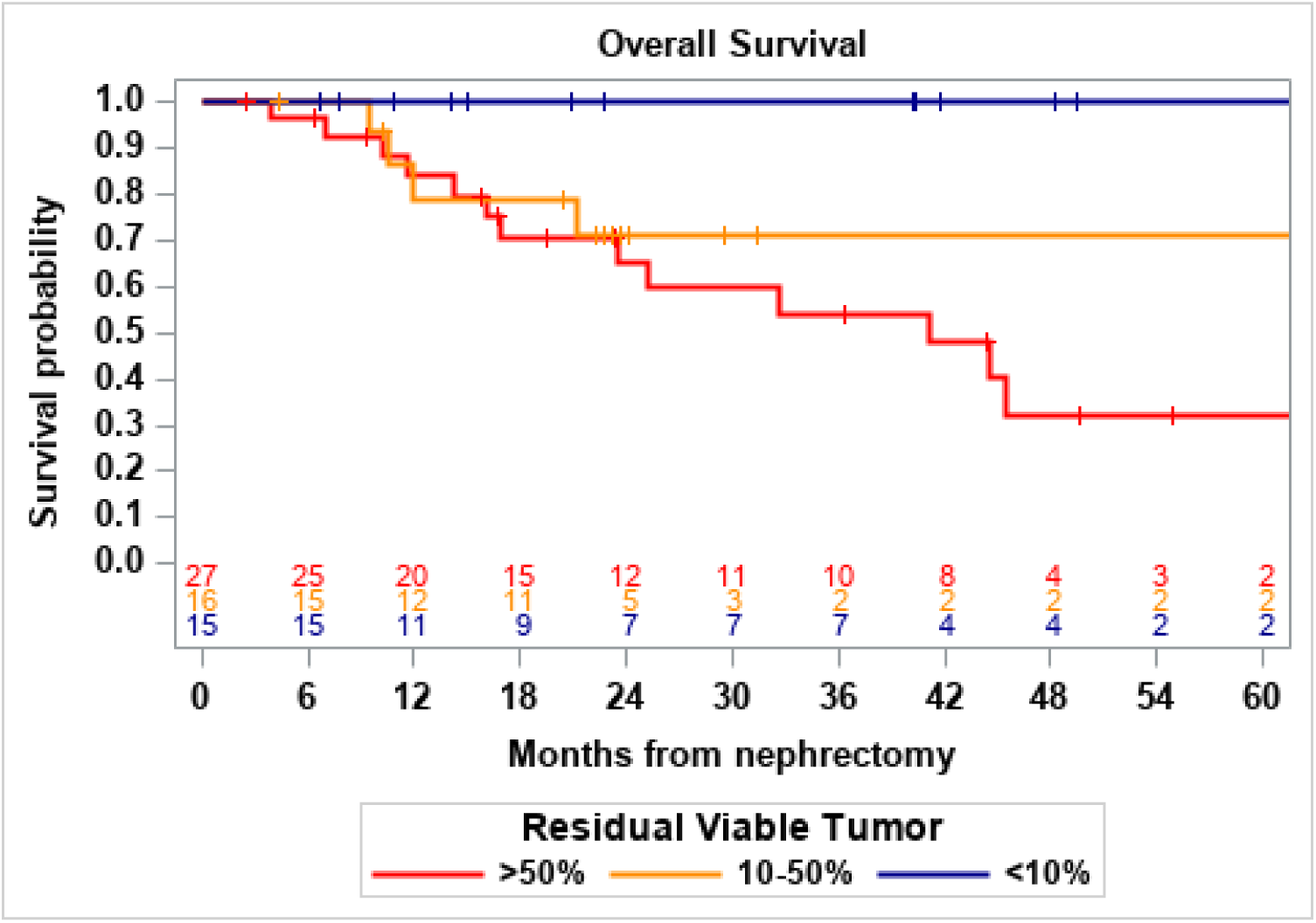
Overall survival by residual viable tumor. In patients with RVT <10%, 10-50% and >50%, 2-year OS was 100%, 76% (95% CI: 43 to 92) and 71% (95% CI: 46 to 86), respectively.

**Table 3.**
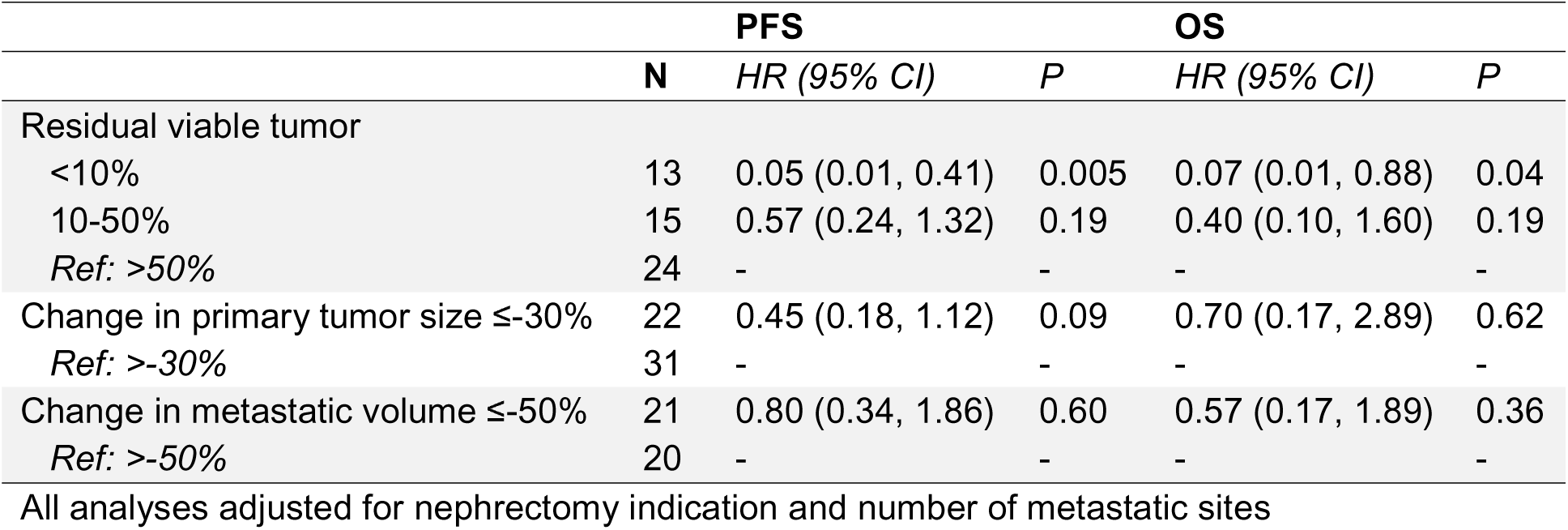
Cox regression analysis of progression-free and overall survival.

In patients undergoing nephrectomy for consolidation, median PFS was 19 months (95% CI: 8, 78), 1-year PFS was 53% (95% CI: 36, 68) and 2-year OS was 81% (95% CI: 62, 91). In patients undergoing CN for palliation, median PFS was 11 months (95% CI: 3, 31), 1-year PFS was 49% (95% CI: 23, 70) and 2-year OS was 60% (95% CI: 32, 80). Kaplan-Meier curves for PFS and OS curves by surgical indication are presented in **Supplementary Figure 1**.

## Discussion

Consolidative nephrectomy after upfront immunotherapy has been retrospectively associated with favorable outcomes in a highly select group of patients with metastatic RCC.^14,19,20^ This study sought to characterize patients undergoing CN at our institution and to identify radiographic and pathologic features associated with disease progression and overall survival.

We retrospectively identified 60 patients who initiated upfront IO therapy at a median of 9 months before CN, which was a similar time frame compared to other modern cohorts which ranged between 4 and 9 months.^11,20^ Our findings also corroborate the favorable OS reported by recent studies of between 4 and 5 years.^19,21^ Though, this study is the first to evaluate distinct clinical features that prognosticate which patients are most likely to benefit from consolidation. In particular, patients with residual viable primary tumor of <10% were associated with favorable PFS and OS. This suggests a potential role for CN in appropriately selected patients after receipt of IO therapy, while residual viable tumor may offer insight into treatment response and prognosis.

Our clinical cohort remains unique in that it is the largest known single center collection of patients undergoing consolidative nephrectomy in the immunotherapy era. Takemura et al. evaluated 40 multi-institutional patients who underwent delayed CN after IO therapy within the International mRCC Database Consortium (IMDC) and reported a similar radiographic response rate of 12/21 patients (57%) with a median OS not reached after a 24 month follow-up period.^11^ Another cohort of 42 prospective mRCC patients who received perioperative nivolumab had an overall response rate of 43% which was similar to the 40% radiographic response rate of our cohort, though they reported a median OS of 44 months which was notably lower than the 78 months in our study.^22^ One explanation for this discrepancy is that they performed a randomized clinical trial evaluating perioperative nivolumab only and did not allow for subsequent lines of therapy. Another non-comparative prospective trial recently reported a median OS of 54.7 months for patients who received an IO combination plus surgery (43 patients).^21^ Yet, none of these studies evaluated radiographic or pathologic variables as predictors for long-term survival outcomes.

While some clinical trials evaluating the efficacy of IO combinations in mRCC have suggested a relationship between radiographic primary tumor shrinkage and survival, this has not previously been evaluated in the post-cytoreductive or consolidative setting.^23,24^ Secondary analyses from CheckMate 214 and KEYNOTE 426 found that a radiologic response < -50% and < -80% of the primary mass within the first 6 months of therapy, respectively, were associated with greater OS in patients without consolidative surgery.^25,26^ In contrast, in our CN cohort, radiographic response based on a cut-off of 30% was not significantly associated with a PFS or OS benefit.^17^ Importantly, we also performed sensitivity analyses evaluating these other radiographic response percentage cut-offs which did not significantly alter this finding.

Studies evaluating primary tumor pathologic response to upfront IO therapy, a potentially key biomarker for guiding adjuvant treatment decisions, have been even more variable due to a lack of consistent assessment criteria. Shapiro et al. reported on a multi-institutional cohort of patients and found that 74 out of 75 patients (98.7%) had residual viable tumor on pathology after IO therapy though the specific criteria and percentages of RVT were not provided.^20^ Older studies have reported on a pT0 rate of between 10 and 14% after IO therapy though they lack the granularity provided by assessment of residual viable tumor.^27,28^ Unfortunately, a standardized procedure for pathologic assessment of post-IO pathologic response has not yet been established for RCC. We therefore reported pathologic response in our study using criteria based on validated guidelines developed for melanoma and found that one-quarter of patients in our cohort had a major pathologic response.^18^ It is worth noting that the routine grossing protocol we utilized in this retrospective cohort included estimation of tumor necrosis quantity, standard tissue representation (≥ 1 section/cm tumor size), and extensive additional sampling if only a small amount of viable tumor was identified on the initial set of slides. Thus, while we feel confident about defining MPR (<10% RVT) in this cohort, further categorization of partial pathologic response (e.g. 10-50%) will require developing a more standardized tumor grossing and sampling method. Still, we hope that this study will set the standard for evaluating pathologic response of the primary tumor for patients undergoing consolidative nephrectomy after immunotherapy.

Due to the current lack of published randomized clinical trials evaluating the efficacy of cytoreductive nephrectomy in the IO era, survival differences based on pathologic response remains unknown. A multi-institutional retrospective study found that downstaging at the time of nephrectomy did not affect 3-year progression-free survival (86.1% vs. 84.2%, p = 0.75) which is similar to findings from a pooled analysis of the National Cancer Database (NCD) which reported no significant decrease in risk of death if there was pathologic downstaging (HR: 0.86, CI: .38 to 1.94, p = 0.70).^19,29^ Though standard pathologic staging often misses nuances of treatment response such as marked central necrosis and tumor viability.^30,31^ Our study is the first to show that a near complete eradication of residual viable tumor may be associated with survival benefit compared to incomplete pathologic responses. This has implications for patient selection, treatment sequencing, and future trial design in this evolving space. Evaluating associations between pathologic MPR and other promising measures of minimal residual disease such as circulating cell-free DNA (cfDNA) and Kidney Injury Molecule-1 (KIM-1) will also be of significant interest in the near future.^32,33^

Our study has several important limitations. First, this is a retrospective study of patients who were selected to undergo consolidative nephrectomy, which was ultimately based on the discretion of the treating physician thus introducing potential selection bias. Second, survival was measured from time of nephrectomy which may result in immortal time bias; though, no patient in our cohort underwent more than two lines of pre-operative systemic therapy and median time from therapy to nephrectomy was consistent with prior reports. Finally, while our cohort size represents one of the largest known series of post-immunotherapy consolidative nephrectomy patients, the sample size limits our ability to draw inferences from differences in our cohort. This in part may explain the lack of statistical significance in some of our Cox regression models despite consistent trends in the hazard ratios supporting the relationship between radiographic shrinkage and survival outcomes. Nevertheless, our study uniquely highlights the importance of including radiographic and pathologic endpoints in future endeavors evaluating potential biomarkers for survival in patients eligible for CN.

## Conclusion

Delayed cytoreductive nephrectomy after immunotherapy is associated with favorable outcomes in select patients with metastatic renal cell carcinoma. Residual viable tumor at time of nephrectomy correlates with prolonged progression-free and overall survival. In contrast, radiographic tumor response did not reliably predict survival in this setting though it did correlate with residual tumor burden. Further prospective studies are needed to validate these findings and refine patient selection for delayed CN after immunotherapy. Upcoming clinical trials should also include radiographic and pathologic endpoints to assess their utility as prognostic biomarkers which could potentially guide post-nephrectomy treatment decisions.

## Data Availability

All data produced in the present study are available upon reasonable request to the authors

## Research support

This work was supported in part by a National Institutes of Health (NIH) / National Cancer Institute cancer center support grant (P30 CA008748) and by the Mazumdar-Shaw Translational Research Initiative in Kidney Cancer. YK is supported by a National Cancer Institute T32 grant (5T32CA082088-25). RRK is supported, in part, by a Department of Defense Early Career Investigator Grant Award (KCRP-AKCI, W81XWH-21-1-0942).

---

**Supplementary Figure 1.**
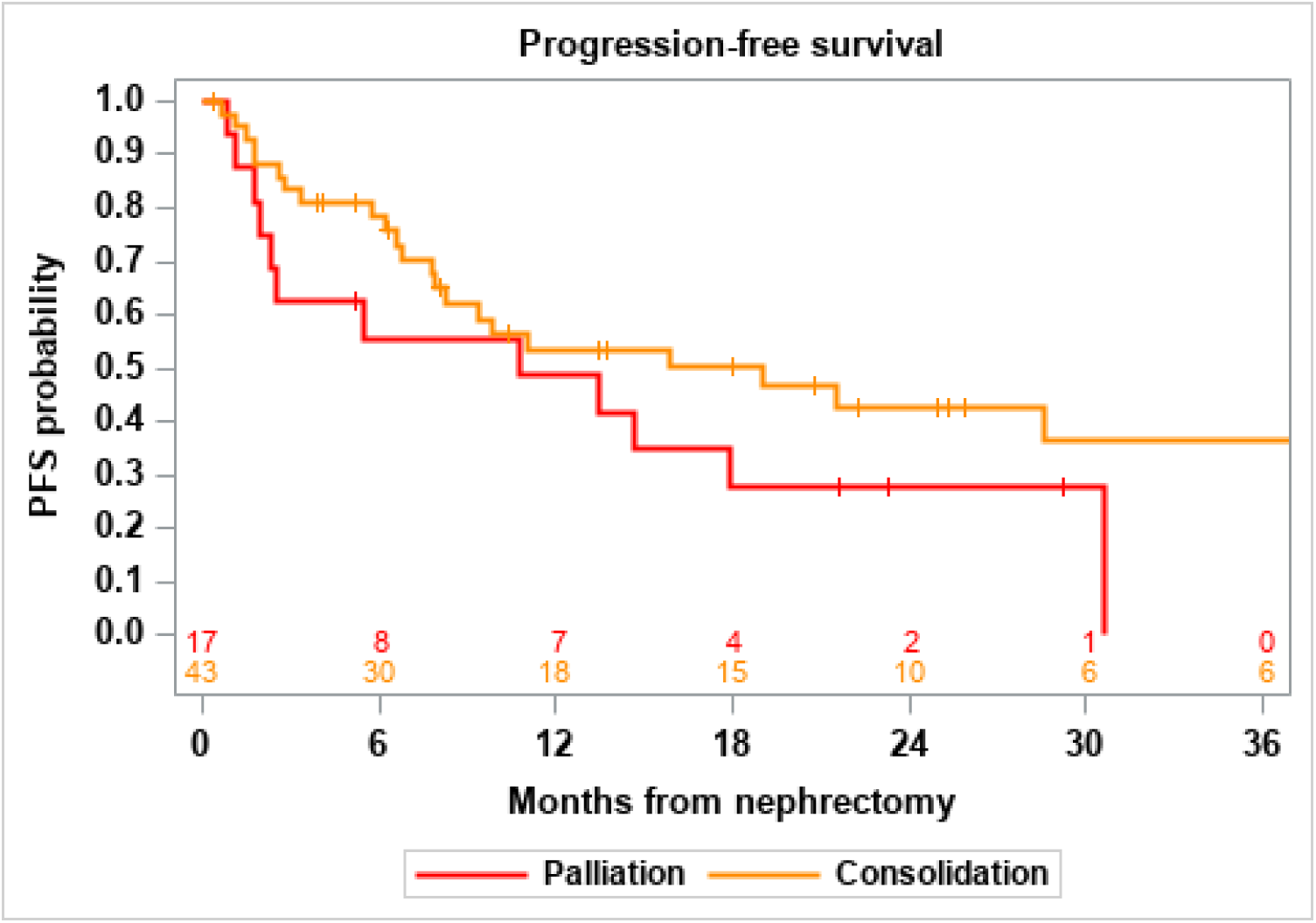
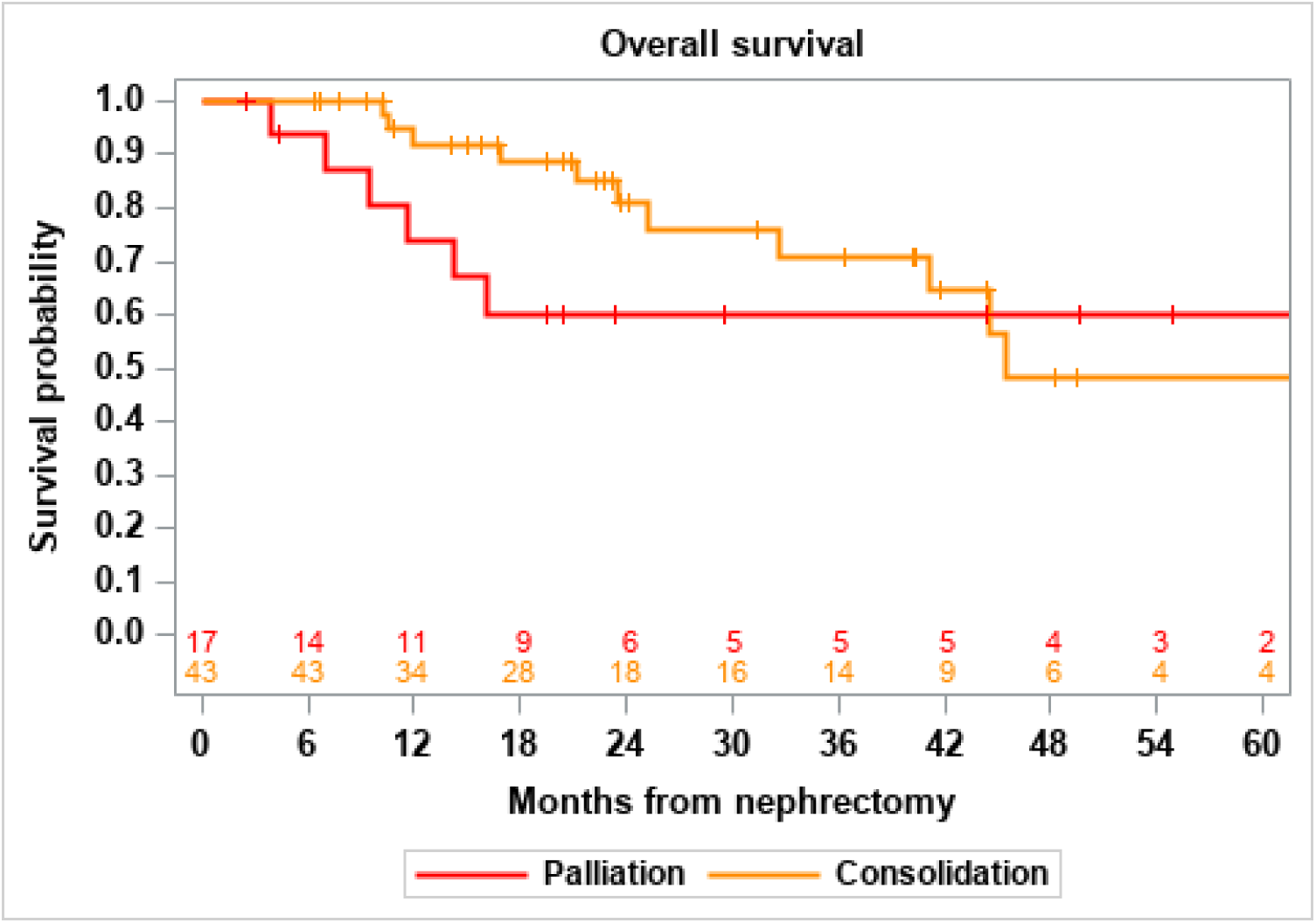
Progression-free and overall survival by nephrectomy indication. In the consolidation patients, median PFS was 19 months (95% CI: 8 to 78), 1-year PFS was 53% (95% CI: 36 to 68) and 2-year OS was 81% (95% CI: 62 to 91). In the palliation patients, median PFS was 11 months (95% CI: 3 to 31), 1-year PFS was 49% (95% CI: 23 to 70) and 2-year OS was 60% (95% CI: 32 to 80).

**Supplementary Table 1.** Post-nephrectomy systemic therapy by nephrectomy indication.

|  | Overall<br>(N=55*) | Consolidation<br>(N=42) | Palliation<br>(N=13) |
| --- | --- | --- | --- |
| Any systemic therapy after nephrectomy | 39 (71%) | 29 (69%) | 10 (77%) |
| Restarted pre-nephrectomy therapy | 31 (56%) | 23 (55%) | 8 (62%) |
| Days from nephrectomy to restart†<br>– median (IQR) | 31 (23, 36) | 31 (23, 36) | 34 (25, 36) |
| Started new therapy (any time after CN) | 18 (33%) | 13 (31%) | 5 (38%) |
| Months from nephrectomy to start of<br>new therapy – median (IQR) | 8.2 (3.8, 12.7) | 9.2 (3.9, 16.8) | 6.2 (2.4, 9.6) |

\*Five patients missing post-nephrectomy systemic therapy information
†All patients restarted within 60 days except one consolidation patient (121 days) and one palliation patient (386 days)

## References

1. Chakiryan NH, Gore LR, Reich RR, et al. Survival Outcomes Associated With Cytoreductive Nephrectomy in Patients With Metastatic Clear Cell Renal Cell Carcinoma. JAMA Netw Open. May 2 2022;5(5):e2212347. doi:10.1001/jamanetworkopen.2022.12347

2. Robert C. Flanigan MD, Sydney E. Salmon, M.D., Brent A. Blumenstein, Ph.D., Scott I. Bearman, M.D., Vivek Roy, M.D., Patrick C. McGrath, M.D., John R. Caton, Jr., M.D., Nikhil Munshi, M.D., and E. David Crawford, M.D. Nephrectomy followed by interferon alfa-2b compared with interferon alfa-2b alone for metastatic renal-cell cancer. The New England Journal of Medicine. October 25, 2001 2001;(345):1655–1659. doi:10.1056/NEJMoa010235

3. Mickisch GH, Garin A, van Poppel H, et al. Radical nephrectomy plus interferon-alfa-based immunotherapy compared with interferon alfa alone in metastatic renal-cell carcinoma: a randomised trial. Lancet. Sep 22 2001;358(9286):966-70. doi:10.1016/s0140-6736(01)06103-7

4. Flanigan RC, Mickisch G, Sylvester R, Tangen C, Van Poppel H, Crawford ED. Cytoreductive nephrectomy in patients with metastatic renal cancer: a combined analysis. J Urol. Mar 2004;171(3):1071–6. doi:10.1097/01.ju.0000110610.61545.ae

5. Mejean A, Ravaud A, Thezenas S, et al. Sunitinib Alone or after Nephrectomy in Metastatic Renal-Cell Carcinoma. N Engl J Med. Aug 2 2018;379(5):417–427. doi:10.1056/NEJMoa1803675

6. Bex A, Mulders P, Jewett M, et al. Comparison of Immediate vs Deferred Cytoreductive Nephrectomy in Patients With Synchronous Metastatic Renal Cell Carcinoma Receiving Sunitinib: The SURTIME Randomized Clinical Trial. JAMA Oncol. Feb 1 2019;5(2):164–170. doi:10.1001/jamaoncol.2018.5543

7. Motzer RJ, Russo P. Cytoreductive Nephrectomy - Patient Selection Is Key. N Engl J Med. Aug 2 2018;379(5):481–482. doi:10.1056/NEJMe1806331

8. Rini BI, Plimack ER, Stus V, et al. Pembrolizumab plus Axitinib versus Sunitinib for Advanced Renal-Cell Carcinoma. N Engl J Med. Mar 21 2019;380(12):1116–1127. doi:10.1056/NEJMoa1816714

9. Motzer R, Alekseev B, Rha SY, et al. Lenvatinib plus Pembrolizumab or Everolimus for Advanced Renal Cell Carcinoma. N Engl J Med. Apr 8 2021;384(14):1289–1300. doi:10.1056/NEJMoa2035716

10. Motzer RJ, Tannir NM, McDermott DF, et al. Nivolumab plus Ipilimumab versus Sunitinib in Advanced Renal-Cell Carcinoma. N Engl J Med. Apr 5 2018;378(14):1277–1290. doi:10.1056/NEJMoa1712126

11. Takemura K, Ernst MS, Navani V, et al. Characterization of Patients with Metastatic Renal Cell Carcinoma Undergoing Deferred, Upfront, or No Cytoreductive Nephrectomy in the Era of Combination Immunotherapy: Results from the International Metastatic Renal Cell Carcinoma Database Consortium. Eur Urol Oncol. Jun 2024;7(3):501-508. doi:10.1016/j.euo.2023.10.002

12. Reese SW, Eismann L, White C, et al. Surgical outcomes of cytoreductive nephrectomy in patients receiving systemic immunotherapy for advanced renal cell carcinoma. Urol Oncol. Feb 2024;42(2):32 e9-32 e16. doi:10.1016/j.urolonc.2023.12.003

13. Fransen van de Putte EE, van den Brink L, Mansour MA, et al. Indications and Outcomes for Deferred Cytoreductive Nephrectomy Following Immune Checkpoint Inhibitor Combination Therapy: Can Systemic Therapy be Withdrawn in Patients with No Evidence of Disease? Eur Urol Open Sci. Sep 2023;55:15–22. doi:10.1016/j.euros.2023.07.002

14. Esagian SM, Karam JA, Msaouel P, Makrakis D. Upfront Versus Deferred Cytoreductive Nephrectomy in Metastatic Renal Cell Carcinoma: A Systematic Review and Individual Patient Data Meta-analysis. Eur Urol Focus. Sep 16 2024;doi:10.1016/j.euf.2024.08.002

15. Heng DY, Xie W, Regan MM, et al. Prognostic factors for overall survival in patients with metastatic renal cell carcinoma treated with vascular endothelial growth factor-targeted agents: results from a large, multicenter study. J Clin Oncol. Dec 1 2009;27(34):5794–9. doi:10.1200/JCO.2008.21.4809

16. Quan H, Li B, Couris CM, et al. Updating and validating the Charlson comorbidity index and score for risk adjustment in hospital discharge abstracts using data from 6 countries. Am J Epidemiol. Mar 15 2011;173(6):676–82. doi:10.1093/aje/kwq433

17. Eisenhauer EA, Therasse P, Bogaerts J, et al. New response evaluation criteria in solid tumours: revised RECIST guideline (version 1.1). Eur J Cancer. Jan 2009;45(2):228–47. doi:10.1016/j.ejca.2008.10.026

18. Tetzlaff MT, Messina JL, Stein JE, et al. Pathological assessment of resection specimens after neoadjuvant therapy for metastatic melanoma. Ann Oncol. Aug 1 2018;29(8):1861–1868. doi:10.1093/annonc/mdy226

19. Singla N, Hutchinson RC, Ghandour RA, et al. Improved survival after cytoreductive nephrectomy for metastatic renal cell carcinoma in the contemporary immunotherapy era: An analysis of the National Cancer Database. Urol Oncol. Jun 2020;38(6):604 e9-604 e17. doi:10.1016/j.urolonc.2020.02.029

20. Shapiro DD, Karam JA, Zemp L, et al. Cytoreductive Nephrectomy Following Immune Checkpoint Inhibitor Therapy Is Safe and Facilitates Treatment-free Intervals. Eur Urol Open Sci. Apr 2023;50:43–46. doi:10.1016/j.euros.2023.01.016

21. Goswami S, Gao J, Basu S, et al. Immune checkpoint inhibitors plus debulking surgery for patients with metastatic renal cell carcinoma: clinical outcomes and immunological correlates of a prospective pilot trial. Nat Commun. Feb 21 2025;16(1):1846. doi:10.1038/s41467-025-57009-z

22. Shen. X.P XM, Wang. JS, Guo.. X. Efficacy of immunotherapy-based immediate cytoreductive nephrectomy vs. deferred cytoreductive nephrectomy in metastatic renal cell carcinoma. European Review for Medical and Pharmacological Sciences. 2023;27:5684–8.

23. Zambrana F, Carril-Ajuria L, Gomez de Liano A, et al. Complete response and renal cell carcinoma in the immunotherapy era: The paradox of good news. Cancer Treat Rev. Sep 2021;99:102239. doi:10.1016/j.ctrv.2021.102239

24. Pieretti AC, Shapiro DD, Westerman ME, et al. Tumor diameter response in patients with metastatic clear cell renal cell carcinoma is associated with overall survival. Urol Oncol. Dec 2021;39(12):837 e9-837 e17. doi:10.1016/j.urolonc.2021.08.022

25. Motzer RJ, Escudier B, McDermott DF, et al. Survival outcomes and independent response assessment with nivolumab plus ipilimumab versus sunitinib in patients with advanced renal cell carcinoma: 42-month follow-up of a randomized phase 3 clinical trial. J Immunother Cancer. Jul 2020;8(2)doi:10.1136/jitc-2020-000891

26. Grünwald V, Choueiri TK, Rini BI, et al. Association between depth of response and overall survival: Exploratory analysis in patients with previously untreated advanced renal cell carcinoma (aRCC) in CheckMate 214. Annals of Oncology. 2019;30:v382–v383. doi:10.1093/annonc/mdz249.046

27. Singla N, Elias R, Ghandour RA, et al. Pathologic response and surgical outcomes in patients undergoing nephrectomy following receipt of immune checkpoint inhibitors for renal cell carcinoma. Urol Oncol. Dec 2019;37(12):924–931. doi:10.1016/j.urolonc.2019.08.012

28. Graafland NM, Szabados B, Tanabalan C, et al. Surgical Safety of Deferred Cytoreductive Nephrectomy Following Pretreatment with Immune Checkpoint Inhibitor-based Dual Combination Therapy. Eur Urol Oncol. Jun 2022;5(3):373–374. doi:10.1016/j.euo.2021.11.004

29. Panian J, Saidian, A., Hakimi, K., Ajmera, A., Anderson, W. J., Barata, P., Berg, S., Signoretti, S., Chang, S. L., D’Andrea, V., George, D., Dzimitrowicz, H., El Zarif, T., Emamekhoo, H., Gross, E., Kilari, D., Lam, E., Lashgari, I., Psutka, S., Rauterkus, G. P., Shabaik, A., Thapa, B., Wang, L., Weise, N., Yim, K., Zhang, T., Derweesh, I., & McKay, R. R. Pathological Outcomes of Patients With Advanced Renal Cell Carcinoma Who Receive Nephrectomy Following Immunotherapy. The Oncologist. 2024;29(10)

30. Atagi Y, Tada K, Kouno R, Minato R, Hashine K. Report of case series: Correlation between pathological and radiological evaluation and clinical course of three cases of metastatic renal cell carcinoma with cytoreductive nephrectomy after combined immuno-oncology therapy. IJU Case Rep. Jul 2024;7(4):341–345. doi:10.1002/iju5.12752

31. Hwang MJ, Brennan PM, Monge BM, et al. Best practices and recommendations for grossing and reporting of post-immunotherapy nephrectomy specimens: a single-institution experience of 70 cases. Diagnostic Histopathology. 2024;30(5):275–281. doi:10.1016/j.mpdhp.2024.02.002

32. Green EA, Li R, Albiges L, et al. Clinical Utility of Cell-free and Circulating Tumor DNA in Kidney and Bladder Cancer: A Critical Review of Current Literature. Eur Urol Oncol. Dec 2021;4(6):893–903. doi:10.1016/j.euo.2021.04.005

33. He S, Liu D, Chen J, et al. Liquid-based kidney injury molecule-1 as a diagnostic and prognostic indicator in renal cell carcinoma: A systematic review and meta-analysis. Int Urol Nephrol. Mar 21 2025;doi:10.1007/s11255-025-04447-9

